# Effectiveness of Covishield vaccine in preventing Covid-19 – A test-negative case-control study

**DOI:** 10.1101/2021.07.19.21260693

**Authors:** Stuti Pramod, Dhanajayan Govindan, Premkumar Ramasubramani, Sitanshu Sekhar Kar, Rakesh Aggarwal, JIPMER vaccine effectiveness study group, Nandhini Manoharan, Palanivel Chinnakali, Mahalakshmy Thulasingam, Sonali Sarkar, Molly Mary Thabah

## Abstract

**Introduction:** This study was aimed at assessing the vaccine effectiveness (VE) of Covishield, which is identical to the AstraZeneca vaccine, in preventing laboratory-confirmed Covid-19.

**Methods:** Using a test-negative case-control design, information on vaccination status of cases with Covid-19 among healthcare workers in our institution in Puducherry, India, and an equal number of matched controls, i.e., positive and negative for SARS-CoV-2 by RT-PCR, was obtained. The cases and controls were matched for age (±3 years) and date of testing (±3 days). The groups were compared using multivariable conditional logistic regression to calculate odds ratios (OR), with adjustment for gender, occupational role, presence of symptoms and presence of a comorbidity condition. Per cent vaccine effectiveness (VE) was calculated as 100 x (1-adjusted odds ratio).

**Results:** Using data from 360 case-control pairs, VE of one dose and of two doses, in providing protection against Covid-19 was 49% (95% CI: 17%-68%) and 54% (27%-71%), respectively. In view of a difference in the proportion of cases and controls who had symptoms, a separate analysis of data from 203 pairs where both the case and the control had symptoms was done, which showed VE of 58% (28%-75%) and 64% (38%-78%) after one dose and two doses, respectively. Among cases with moderately severe disease that required oxygen therapy, VE following any number of vaccine doses was 95% (44%-100%).

**Conclusion:** Covishield vaccine protected significantly against Covid-19, with the protection after two doses being slightly higher than after one dose, and a particularly high protection rate against severe forms of disease.

## Introduction

Vaccination is an important measure for preventing Covid-19. India started vaccination against Covid-19 on 16^th^ January 2021 in a phased manner, prioritising first the health care workers (HCWs) and other frontline workers, extending it to those over 60 years of age and those aged 45-60 years with comorbidities, then to all those aged 45 years and above, and finally to all adults.^1^ Two vaccines were authorized by the Indian drug regulator for emergency use, namely Covishield (a recombinant, replication-deficient chimpanzee adenovirus vector that encodes SARS-CoV-2 spike glycoprotein) and Covaxin (inactivated whole virions grown in Vero cells). Covishield, which is identical to the Oxford-AstraZeneca (ChAdOx1 nCoV-19) vaccine in composition and immunogenicity,^2^ has accounted for nearly 88% of all doses in the country to date, and has been the sole vaccine used in some areas, including our city.^3^ In pooled data from four trials, this vaccine had a protective efficacy of 67% (95% confidence interval [CI]: 57%-74%) for preventing symptomatic and laboratory-proven Covid-19 and of nearly 100% (72-100%) for preventing hospitalization and severe infection, beginning 21 days after the second dose.^4^

Since clinical trials include selected individuals, it is important for the protection offered by any vaccine to be studied in real-world settings. A commonly used method for evaluating population-level effectiveness of covid vaccines has been to assess their effect in preventing infection, which is defined as detection of viral RNA or antigen in a respiratory specimen collected from a person, after a specified period has elapsed after receipt of all recommended doses, using a case-control design.^5^

Our study was aimed at determining the effectiveness of the Covishield vaccine in preventing laboratory confirmed Covid-19, separately for those who had received a single dose and for those who had received two doses of this vaccine.

## Methods

We designed a test-negative case-control study in our institution, a large teaching hospital, located in the Puducherry district in Southern India, on the East coast, with nearly 8,700 healthcare workers (HCWs). Vaccination with Covishield had started in our institution on 16^th^ January 2021, though some of our HCWs could receive the other vaccine by travelling to other areas. Assuming a vaccination coverage of 50%, effectiveness of 70%, and a matched case-control design, the sample size was calculated to be 346 case-control pairs. The study was approved by our institution’s ethics committee and all subjects provided an oral informed consent.

A case was defined as a HCW in our institution who had tested positive for active SARS-CoV-2 infection using RT-PCR during March 1-May 31, 2021. Students, whether medical, nursing and other allied health science, were not considered as HCWs. All the consecutive cases identified were contacted, and were enrolled if they agreed. In persons who had more than one positive test result, the date of first positive report was used.

A control was a HCW aged within 3 years of the particular case, and who had tested negative for SARS-CoV-2 by RT-PCR within 3 days of the particular case testing positive. Persons with negative SARS-CoV-2 antigen test alone were not included.

Data on vaccination status, type of vaccine, test positivity, presence of symptoms and comorbidities were collected using a telephonically administered questionnaire, captured using EpiCollect5 application and analysed using STATA V14.0. To calculate vaccine effectiveness (VE), the protective effect was taken as appearing 21 days after vaccination if only one dose had been administered and 14 days after the second dose if two doses had been administered. Matched-pair analysis was done, and univariate and multivariable conditional logistic regression was done to calculate unadjusted and adjusted odds ratios. Factors used for adjustment included gender, occupational role, presence of any comorbidity and presence of symptoms at the time of RT-PCR testing. Percent VE was calculated as 100*(1-odds ratio). A subgroup analysis was done for cases with moderately severe disease and their matched controls, to look specifically at the VE against such disease.

In addition, a separate analysis was done for the pairs where both the case and the respective control were symptomatic.

## Results

Our database showed that around 2200 tests had been performed in our HCWs from March to May 2021. Of these, 795 were positive. To recruit 360 cases and 360 matched controls, we contacted 547 test-positive HCWs (65.8% response) and 963 test-negative HCWs (37.3% response), respectively (Table 1). Their median (interquartile range) age was 34 (28-43) and 33 (28-42) years, respectively. The distribution of gender and comorbidities was comparable between cases and controls. Among cases, 15% (n=54) had one or more comorbidities, of which the most common were hypertension (n=29; 8.1%) and diabetes mellitus (n=25; 7.0%). Most of the cases had mild disease requiring only home isolation (n=350; 97.2%).

**Table 1:** Characteristics of cases with Covid-19 and controls matched for age and day of onset of illness, March–May 2021, Puducherry, India

| Characteristic | Cases<br>(n= 360) |  | Matched controls<br>(n=360) |  | P value |
| --- | --- | --- | --- | --- | --- |
| **Age categories, number (%)** |  |  |  |  |  |
| ≤ 29 | 108 | (30.0) | 115 | (31.9) |  |
| 30 to 39 | 130 | (36.1) | 135 | (37.5) |  |
| 40 to 49 | 84 | (23.3) | 75 | (20.8) |  |
| ≥ 50 | 38 | (10.6) | 35 | (9.7) |  |
| **Gender, number (%)** |  |  |  |  |  |
| Men | 178 | (49.5) | 181 | (50.3) | 0.819* |
| Women | 182 | (50.5) | 179 | (49.7) |  |
| **Occupational role<sup>†</sup>** |  |  |  |  |  |
| Doctors | 49 | (13.6) | 71 | (19.7) | <0.001^ |
| Nursing staff | 138 | (38.3) | 173 | (48.1) |  |
| Paramedical or Support staff | 136 | (37.8) | 84 | (23.3) |  |
| Administrative staff | 37 | (10.3) | 32 | (8.9) |  |
| **Vaccination status at enrollment** |  |  |  |  |  |
| Unvaccinated | 148 | (41.1) | 57 | (15.8) |  |
| Received one dose | 114 | (31.7) | 140 | (38.9) |  |
| Received two doses | 98 | (27.2) | 163 | (45.3) |  |
| **Comorbidity<sup>§</sup>** |  |  |  |  |  |
| Yes | 54 | (15.0) | 42 | (11.7) | 0.188* |
| No | 306 | (85.0) | 318 | (88.3) |  |
| **Covid symptoms at the time of RT-PCR test** |  |  |  |  |  |
| Yes | 343 | (95.3) | 212 | (58.9) | <0.001* |
| No | 17 | (4.7) | 148 | (41.1) |  |
| **Time of RT-PCR test** |  |  |  |  |  |
| March 2021 | 19 | (5.3) | 19 | (5.3) |  |
| April 2021 | 95 | (26.4) | 93 | (25.8) |  |
| May 2021 | 246 | (68.3) | 248 | (68.9) |  |

\* McNemar test, ^Chi squared test
§ Chronic lung disease, malignancy, heart disease, chronic liver disease, chronic renal disease, diabetes, hypertension or immunocompromised state
† Paramedical or support staff include laboratory technicians, operation theatre technicians, radiographers, ward attendants, drivers, security staff, sanitation worker

All the vaccinated subjects among both cases and controls had received Covishield, and none had received another Covid-19 vaccine. Considering the onset of protection as 21 days after the first dose or 14 days after the second dose, and after adjustment for gender, occupational role, comorbidity, symptomatology, the effectiveness of one dose and two doses, in protecting against Covid-19 was found to be 49% (95% CI: 17%-68%) and 54% (27%-71%), respectively (Table 2).

**Table 2:**
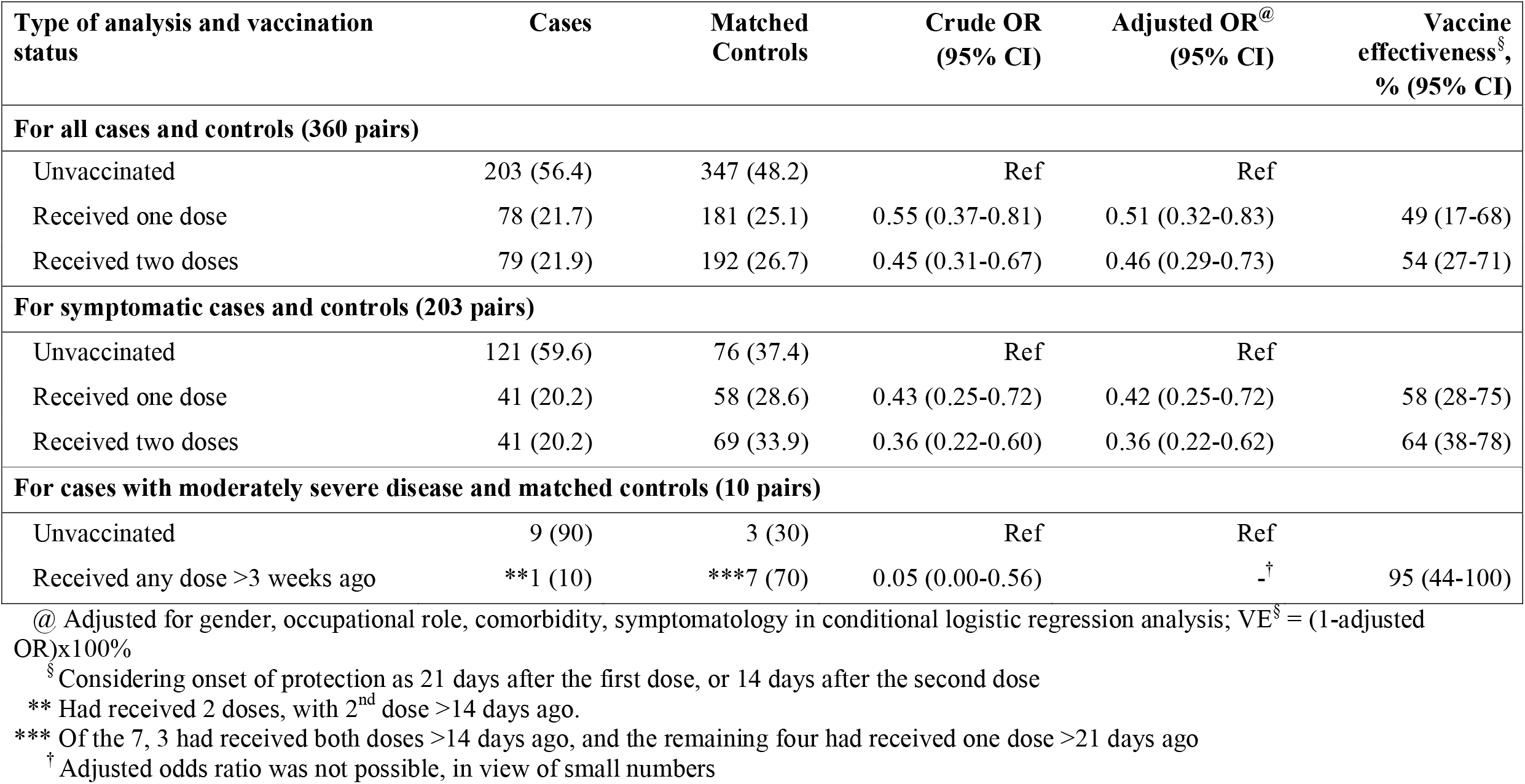
Comparison of vaccination status between cases and matched controls, and adjusted vaccine effectiveness against Covid-19

In the analysis of data from 203 case-control pairs where both cases and controls had symptoms, after adjustment for various factors, the VE associated with one dose and two doses was 58% (28%-75%) and 64% (38%-78%), respectively.

In a subgroup analysis of the 10 cases (2.8%) who had a moderately severe disease that required oxygen therapy, nine were found to be unvaccinated. By contrast, only 3 of their 10 matched controls were unvaccinated (p=0.019, Fisher’s exact test; VE = 95% [44%-100%]).

## Discussion

Our data show that vaccination was associated with a reduction in the risk of Covid-19 and, in particular, of moderately severe disease needing hospital care, among HCWs in our institution.

Another test-negative case-control study from Vellore, India showed VE among HCWs who had received two doses of a Covid-19 vaccine to be 65% (95% CI: 61-68), ^6^ which was somewhat higher than that in our study. This difference in VE can be related to many factors, such as differences in the prevalent virus strains, overall disease endemicity and vaccination coverage. Though Vellore is situated fairly close to our city, the study there included cases that occurred between mid-January 2021 and April 2021, whereas our study included those from March to May 2021. It is well known that the number of cases with the delta variant (B.1.617.2), a variant of concern, of SARS-CoV-2 surged in India during March to May 2021. Thus, during the period of our study, over 70% of cases in Puducherry were caused by this variant.^7^ Further, a higher overall disease rate in our area and a lower vaccine coverage rate among our HCWs than those in the Vellore study could also explain the observed difference. The fact that some (nearly 7%) of subjects in the Vellore study and none in our study had received Covaxin, the other vaccine available in India, is unlikely to have made a difference.

There has been only one other published report from India on the protection afforded by Covid-19 vaccines.^8^ It showed that the proportion of those who had received one or two doses of a Covid-19 vaccine had a lower risk of having Covid-19; however, since no data were provided on the time interval between vaccine doses and disease, a formal calculation of VE was not possible.

Several studies from other parts of the world have assessed the VE of the AstraZeneca vaccine, which Covishield is identical to. A cohort study conducted in Chile between February 2021 through May 2021 showed VE of 65.9% (95% CI: 65.2-66.6) among the fully immunized.^9^ In a cohort study conducted in Scotland, the vaccine effect for this vaccine was 88% (95% CI: 75-94) between December 2020 to February 2021.^10^ In the United Kingdom, VE against B.1.617.2 variant was estimated to be 32.9 (95% CI: 19.3-44.3) after only the first dose and 59.8% (95% CI: 28.9-77.3) after two doses of this vaccine.^11^ Real-life data for Covid-19 vaccines based on mRNA platform have also shown similarly high VE.

The test-negative case-control study design is efficient and eliminates bias stemming from differences in healthcare-seeking behaviour and community-level variations in vaccine access and disease risk.^12^ However, since we found a difference in the frequency of symptoms among our cases and controls, we undertook an additional analysis to control for this factor. In this analysis restricted to only those case-control pairs where both the case and the control had symptoms, the VE estimates for one as well as two doses were somewhat better than those in our primary analysis. This may indicate that the vaccine may in fact have a better efficacy than suggested in our initial analysis above.

Importantly, our study showed that the VE of Covishield against moderately-severe disease was much higher than that against disease of any severity. This is an important finding since the primary aim of Covid-19 vaccination is to prevent serious disease needing hospitalization so that healthcare facilities are not overwhelmed and lives are not lost. It has been difficult to reliably assess the efficacy of Covid-19 vaccines in clinical trials against moderate to severe disease because of relative infrequency of the latter. The case-control design that we used, despite its several limitations, has the advantage of permitting assessment of association of intervention with disease even when only a few cases are available, and thus allowed us to detect this effect. Though our analysis did show a statistically significant protection against moderately severe disease after Covishield, the confidence intervals of the estimate are relatively broad and further data on this association may be needed to improve our confidence in this observation.

Our study has two key limitations. First, the study design used relies heavily on reporting for RT-PCR testing. Thus, it may overestimate the benefit of vaccination if the vaccinated HCW, whether asymptomatic or having symptoms suggestive of Covid-19, were to believe that they were unlikely to have Covid-19 and decide not to report for testing. Second, since genomic sequencing of SARS-CoV-2 had not been done in our study participants, we were unable to assess the VE separately for the ancestral strain and variant strains of the virus.

In conclusion, our data show that Covishield vaccination, either as one dose or in a 2-dose schedule, was effective in halving the frequency of Covid-19 disease among HCWs in a period when B.1.617.2 strain of SARS-CoV-2 was the dominant strain circulating in our area, and had an even greater effect on preventing a more severe and clinically relevant form of this disease.

## Data Availability

As per the institute’s ethics guidelines, the ethics committee reserves the rights on the datasets which protects the privacy of research data for up to three years after completion of the studies. Therefore, the dataset cannot be shared at this point in time. In case of any further queries or clarifications regarding the institute’s ethics guidelines on sharing of research data, the committee may be contacted through their official The data may be accessed from the JIPMER Ethics Committee (contact) after three years of completion of the project, for those who meet institute’s criteria for access to confidential data.

## Acknowledgement

We thank the members of our survey team, data management group and the study participants for their help.

